# Chest Computed Tomography for the Diagnosis of Patients with Coronavirus Disease 2019 (COVID-19): A Rapid Review and Meta-Analysis

**DOI:** 10.1101/2020.04.14.20064733

**Authors:** Meng Lv, Mengshu Wang, Nan Yang, Xufei Luo, Wei Li, Xin Chen, Yunlan Liu, Mengjuan Ren, Xianzhuo Zhang, Ling Wang, Yanfang Ma, Junqiang Lei, Toshio Fukuoka, Hyeong Sik Ahn, Myeong Soo Lee, Zhengxiu Luo, Yaolong Chen, Enmei Liu, Jinhui Tian, Xiaohui Wang, on behalf of COVID-19 evidence and recommendations working group

## Abstract

**Background:** The outbreak of the coronavirus disease 2019 (COVID-19) has had a massive impact on the whole world. Computed tomography (CT) has been widely used in the diagnosis of this novel pneumonia. This study aims to understand the role of CT for the diagnosis and the main imaging manifestations of patients with COVID-19.

**Methods:** We conducted a rapid review and meta-analysis on studies about the use of chest CT for the diagnosis of COVID-19. We comprehensively searched databases and preprint servers on chest CT for patients with COVID-19 between 1 January 2020 and 31 March 2020. The primary outcome was the sensitivity of chest CT imaging. We also conducted subgroup analyses and evaluated the quality of evidence using the Grading of Recommendations Assessment, Development and Evaluation (GRADE) approach.

**Results:** A total of 104 studies with 5694 patients were included. Using RT-PCR results as reference, a meta-analysis based on 64 studies estimated the sensitivity of chest CT imaging in COVID-19 was 99% (95% CI, 0.97-1.00). If case reports were excluded, the sensitivity in case series was 96% (95% CI, 0.93-0.99). The sensitivity of CT scan in confirmed patients under 18 years old was only 66% (95% CI, 0.11-1.00). The most common imaging manifestation was ground-glass opacities (GGO) which was found in 75% (95% CI, 0.68-0.82) of the patients. The pooled probability of bilateral involvement was 84% (95% CI, 0.81-0.88). The most commonly involved lobes were the right lower lobe (84%, 95% CI, 0.78-0.90) and left lower lobe (81%, 95% CI, 0.74-0.87). The quality of evidence was low across all outcomes.

**Conclusions:** In conclusion, this meta-analysis indicated that chest CT scan had a high sensitivity in diagnosis of patients with COVID-19. Therefore, CT can potentially be used to assist in the diagnosis of COVID-19.

## Background

In early January 2020, a disease caused by a novel coronavirus rapidly spread and across the whole world. The disease was later named as Coronavirus disease 2019 (COVID-19). On 11 March 2020, COVID-19 was declared by the WHO a pandemic (1). As of 12 April 2020, the World Health Organization (WHO) has reported 1,614,951 confirmed cases across more than 200 countries (2).

COVID-19 is a respiratory illness that can spread from human to human. Patients with the disease have mild to severe respiratory illness with symptoms such as fever, cough, dyspnea, as well as other non-specific symptoms including, fatigue, myalgia, and headache (3-5). Based on current knowledge, the median basic reproductive number (R_0_) value of COVID-19 is 5.7 (95%CI 3.8-8.9) (6), which means that COVID-19 is highly contagious.

COVID-19 is mainly diagnosed by viral nucleic acid test, immunological detection, and radiological examination. However, the sensitivity of the nucleic acid test may be as low as 50% (7), and some diagnoses may be missed. As a respiratory disease, imaging detection plays an important role in the diagnosis of COVID-19. On one hand, when COVID-19 cannot be diagnosed by nucleic acid, CT can be used as an auxiliary diagnostic method; on the other hand, CT can show lesions and also plays an important role in patient follow-up. Since February 2020, several case-control studies (8,9), case series (10,11), and case-report (12,13) of CT diagnosis of COVID-19 have been published. However, there is no systematic review and meta-analysis to find out the performance of chest CT in the diagnosis of COVID-19. We therefore conducted this study to estimate the sensitivity of chest CT and the probability of imaging findings in cases with COVID-19 to guide the diagnosis of COVID-19.

## Methods

### Search strategy

We searched Medline (via PubMed), Embase, Cochrane library, Web of Science, China Biology Medicine disc (CBM), China National Knowledge Infrastructure (CNKI) between 1 January 2020 and 31 March 2020, using terms with (“2019-novel coronavirus” OR “Novel CoV” OR “2019-nCoV” OR “2019-CoV” OR “Wuhan-Cov” OR “Wuhan Coronavirus” OR “Wuhan seafood market pneumonia virus” OR COVID-19 OR SARS-CoV-2 OR “novel coronavirus pneumonia”) AND (“computed tomography” OR “radiograph*” OR imagine*) The details of the search strategy can be found in the ***Supplementary Material 1***. We also searched Google Scholar and the preprint servers, including SSRN (https://www.ssrn.com/index.cfm/en/), medRxiv (https://www.medrxiv.org/) and bioRxiv (https://www.biorxiv.org/), as well as reference lists of the identified articles, to find additional studies. This systematic review and meta-analysis followed the Preferred Reporting Items for Systematic Reviews and Meta-analysis (PRISMA) statements checklist (14).

### Inclusion and exclusion criteria

In this study, we included records that focused on chest CT imaging for patients with COVID-19 published or posted in English or Chinese. We included original studies fulfilling the following criteria:1) the study topic is related to chest CT manifestations during COVID-19 diagnosis, 2) the participants are children or adults who had an eventual confirmed diagnosis of COVID-19 by RT-PCR testing, and 3) study design is case series and case report. We excluded studies with insufficient data and no response from the author, and studies for which we could not access the full text.

### Selection of studies

Two trained researchers (M Lv and M Wang) screened titles, abstracts, and the full texts of the identified studies independently using Endnote X9 software. Discrepancies were resolved through consultation with a third researcher. We first conducted a pretest with a small sample before the full screening, followed by discussion, to improve the consistency between the reviewers. All reasons for excluding ineligible studies were documented, and the study selection process was documented using a PRISMA flow chart.

### Data extraction

Eight researchers (N Yang, X Luo, W Li, X Chen, Y Liu, M Ren, X Zhang and L Wang) were divided into four groups to extract the data and collect the following information for each study: basic information (title, first author, country or region of participants, date of publication/posting, journal, and study type), patient information (sample size, female/male ratio, adult/children ratio, age range, mean age), outcome information (primary outcome: sensitivity of chest CT imaging using reverse transcription polymerase chain reaction (RT-PCR) results as reference; other outcomes, including probability of bilateral or unilateral pneumonia, ground-glass opacities (GGO) and consolidation, number of lobes affected, location of lobe involvement, rounded morphology, linear opacities, crazy-paving pattern, air bronchogram, interlobular septum thickening, pleural thickening, halo sign, reverse halo sign, pleural effusion and lymphadenopathy).

### Risk of bias assessment

Two researchers assessed the methodological quality of cases series and case reports using the revised checklist form of Murad *et al* (15). The Murad *et al* checklist contains a total of eight items, grouped into four domains (selection, ascertainment, causality and reporting). A pretest was performed before the formal assessment to ensure that the reviewers understood the criteria and process of evaluation consistently. Disagreements were solved by discussion or consultation with a third researcher.

### Data synthesis

We performed a meta-analysis using STATA 15.1. We present data from eligible studies in an evidence table and using descriptive statistics. The percentages of the sensitivity of CT examination and the probability of imaging manifestations in patients with COVID-19 were computed using the *metaprop* command (Stata) for the meta-analysis of proportions. *metaprop* allows the inclusion of studies with proportions equal to 0 or 100% and avoids CIs surpassing the 0 to 1 range, where normal approximation procedures often break down. It achieves this by using the binomial distribution to model within-study variability or by allowing Freeman-Tukey double arcsine transformation to stabilize the variances.

We generated a forest plot to show the individual and pooled probabilities of positive initial CT examination, their 95%□CI and study weights. We conducted subgroup analyses based on case series, and children (≤18).

### Quality of the evidence assessment

The quality of evidence for each outcome was assessed using the the Grading of Recommendations Assessment, Development and Evaluation (GRADE) approach (16). The criteria mainly considered included study methodological quality, directness of the evidence, heterogeneity of data, precision of effect estimates, and risk of publication bias (17-21). The quality of evidence for each outcome was graded as high, moderate, low, or very low.

As COVID-19 is a public health emergency of international concern and the situation is evolving rapidly, our study was not registered in order to speed up the process.

## Results

### Study selection and characteristics

The literature search retrieved 545 records. After the removal of 441 studies not meeting the inclusion criteria, 104 studies with a total of 5694 participants were eligible for inclusion (*Figure 1*) (*List of included studies see Supplementary material 2*). The studies were published between 4 February 2020 and 31 March 2020.

**Figure 1.**
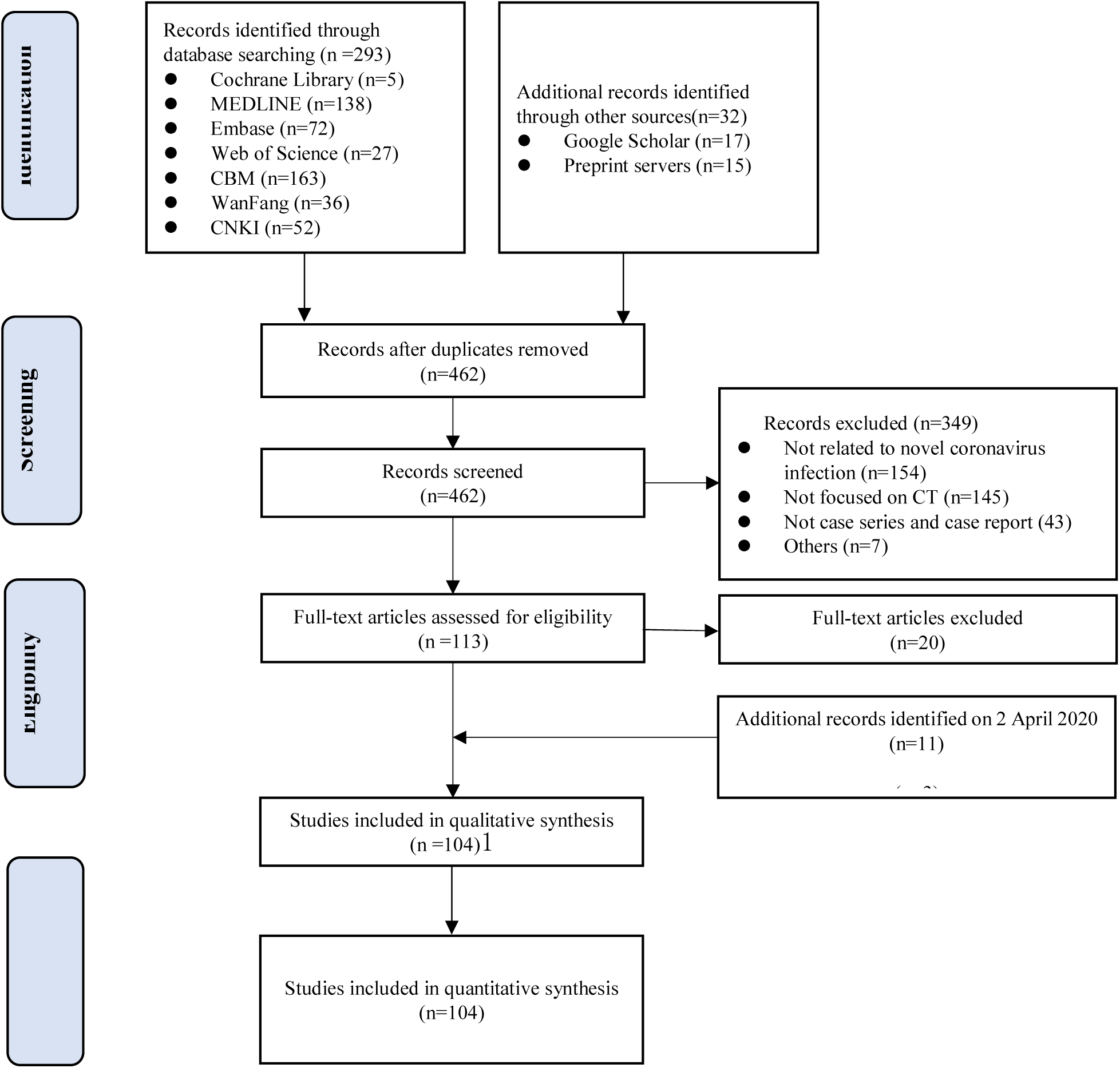
Flowchart of study selection process and results

### Study characteristics and risk of bias

Of the 104 included studies, 83 were case series and 21 were case reports. Ninety-five studies included cases from China, and one each from Germany, Korea, Italy and the cruise ship “Diamond Princess”. The characteristics of included studies were summarized in *Supplementary Material 3*. In 54 of the 104 case series and case reports the overall score was below 50%, indicating a high risk of bias (*see Supplementary material 4*).

### Performance of chest CT in diagnosing COVID-19

The result of meta-analysis showed that using RT-PCR results as reference, the pooled sensitivity of chest CT imaging of sixty-four studies with 3243 COVID-19 patients was 99% (95% CI, 0.97-1.00, *I*^*2*^=85.00%). The quality of evidence was low (*Figure 2*).

**Figure 2.**
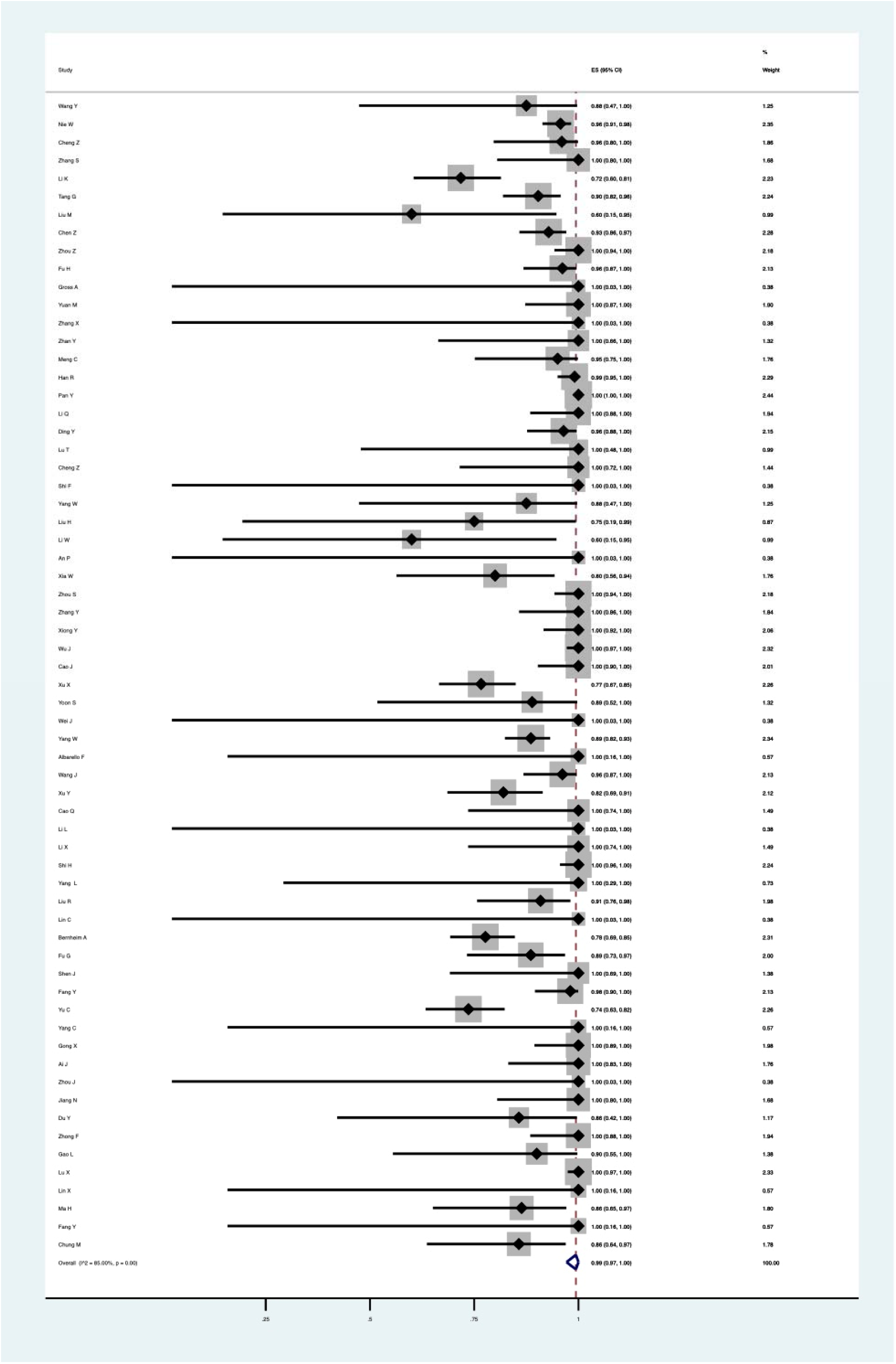
Meta-analyses of the sensitivity of chest CT scan in COVID-19

### Subgroup analyses

In a subgroup analysis of 47 case series (excluding the case reports) the sensitivity of chest CT imaging was 96% (95% CI, 0.93-0.99, *I*^*2*^=88.70%). The quality of evidence was low (*Figure 3*). The subgroup analysis of sensitivity of chest CT imaging in children based on seven studies was 66% (95% CI, 0.11-1.00, *I*^*2*^=96.74%) (*Figure 4*).

**Figure 3.**
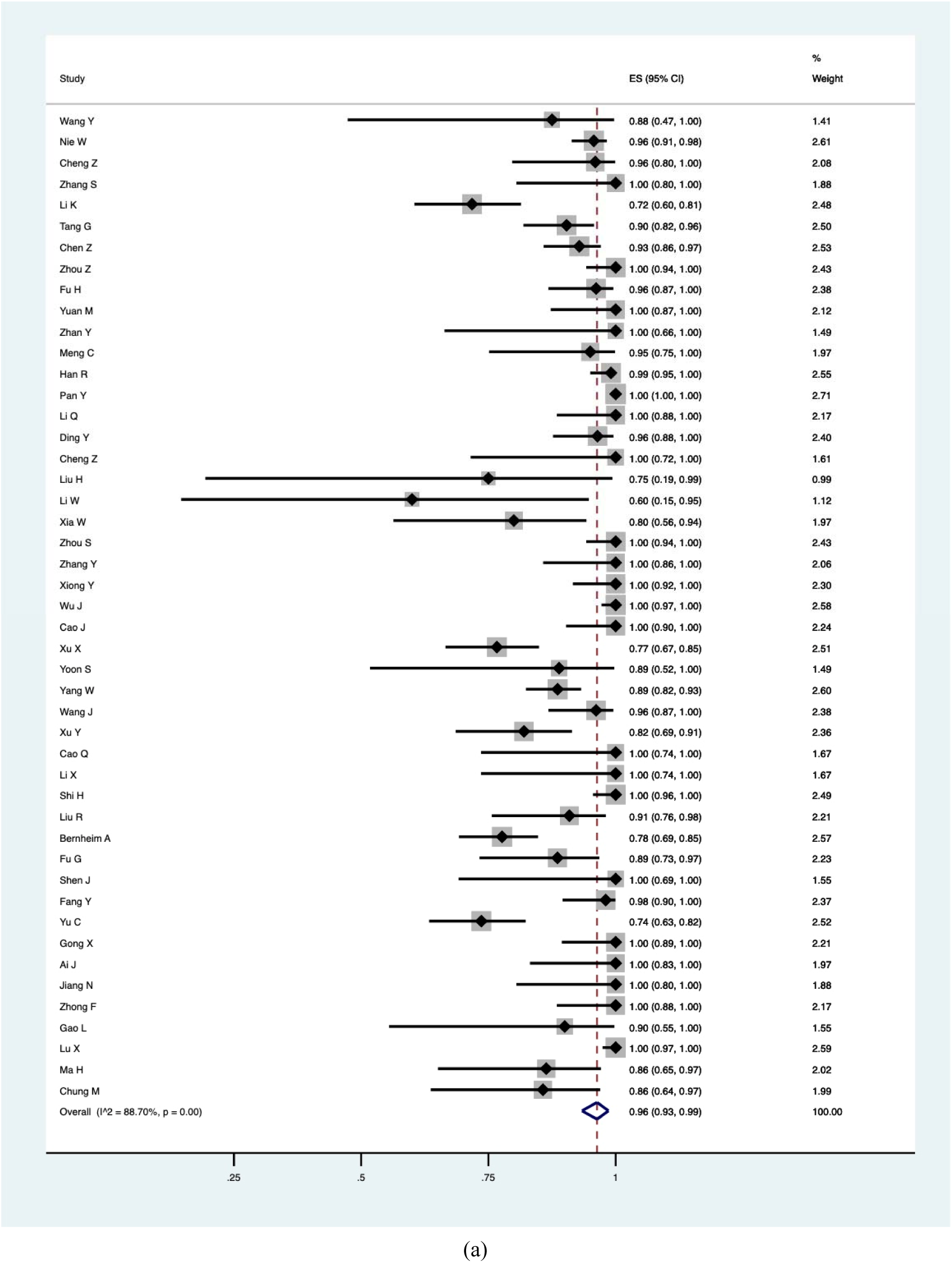
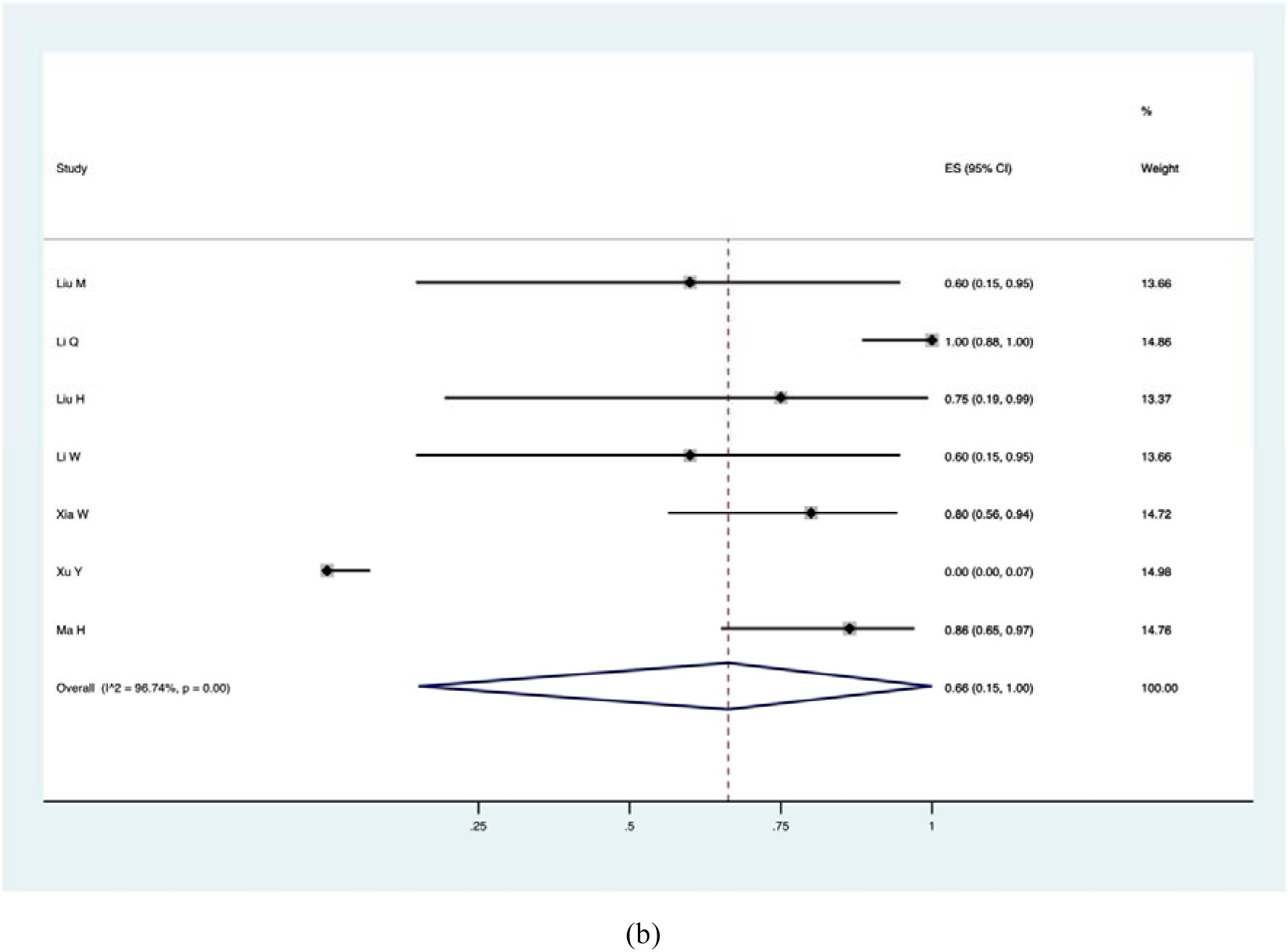
Subgroup analyses of the sensitivity of chest CT scan: case series (panel a); and children (panel b).

**Figure 4.**
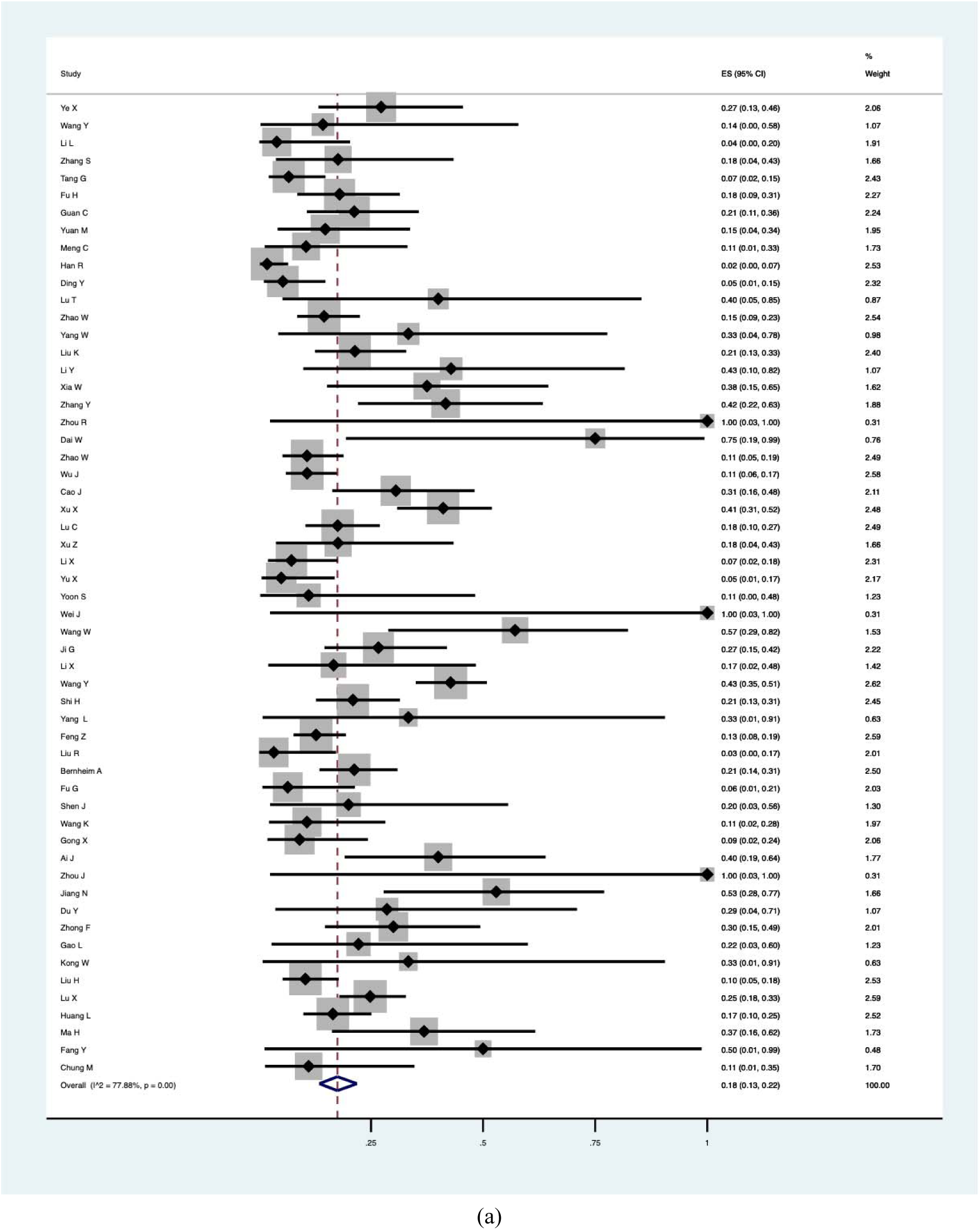
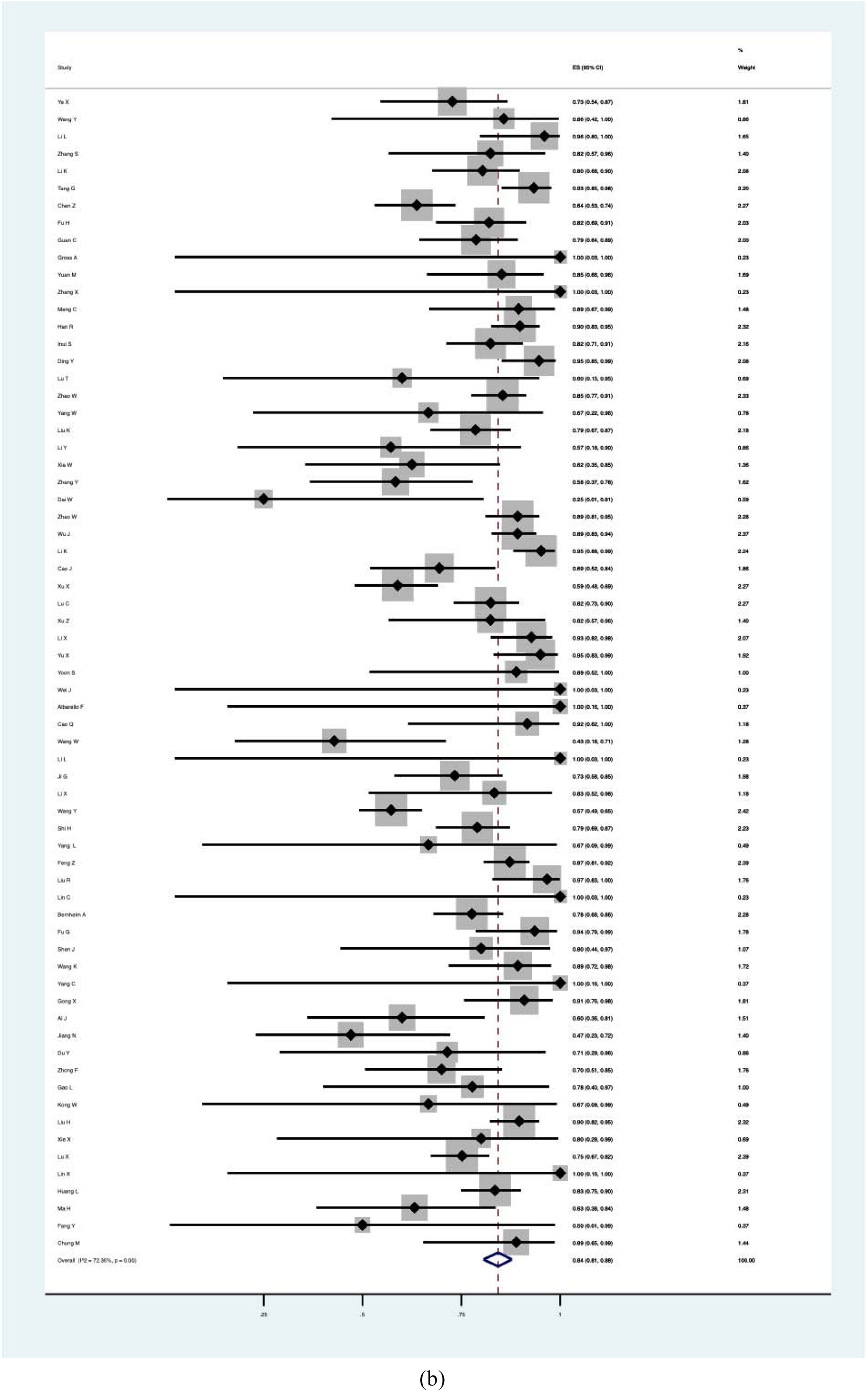
Meta-analyses of the probability of unilateral (panel a) and bilateral (panel b) involvement

### Probability of unilateral/bilateral involvement

Fifty-six studies reported the probability of unilateral involvement. Our meta-analysis showed that the pooled probability of unilateral involvement was 18% (95% CI, 0.13-0.22, *I*^*2*^=77.88%). Sixty-seven studies reported the probability of bilateral involvement, and the pooled probability was 84% (95% CI, 0.81-0.88, *I*^*2*^=72.35%). The quality of evidence for both outcomes was low (*Figure 4*).

### Probability of lesion density

Seventy-five studies reported the proportion of the patients with GGO. The meta-analysis showed that the probability of GGO was 75% (95% CI, 0.68-0.82, *I*^*2*^= 94.32%). The pooled probability of consolidation, based on a meta-analysis of 42 studies, was 34% (95% CI, 0.23-0.45, *I*^*2*^= 95.80%). Fifty-one studies reported the probability of GGO with consolidation, which was estimated 48% (95% CI, 0.40-0.56, *I*^*2*^= 88.98%) in the meta-analysis. The quality of evidence for all three outcomes was low (*Figure 5*).

**Figure 5.**
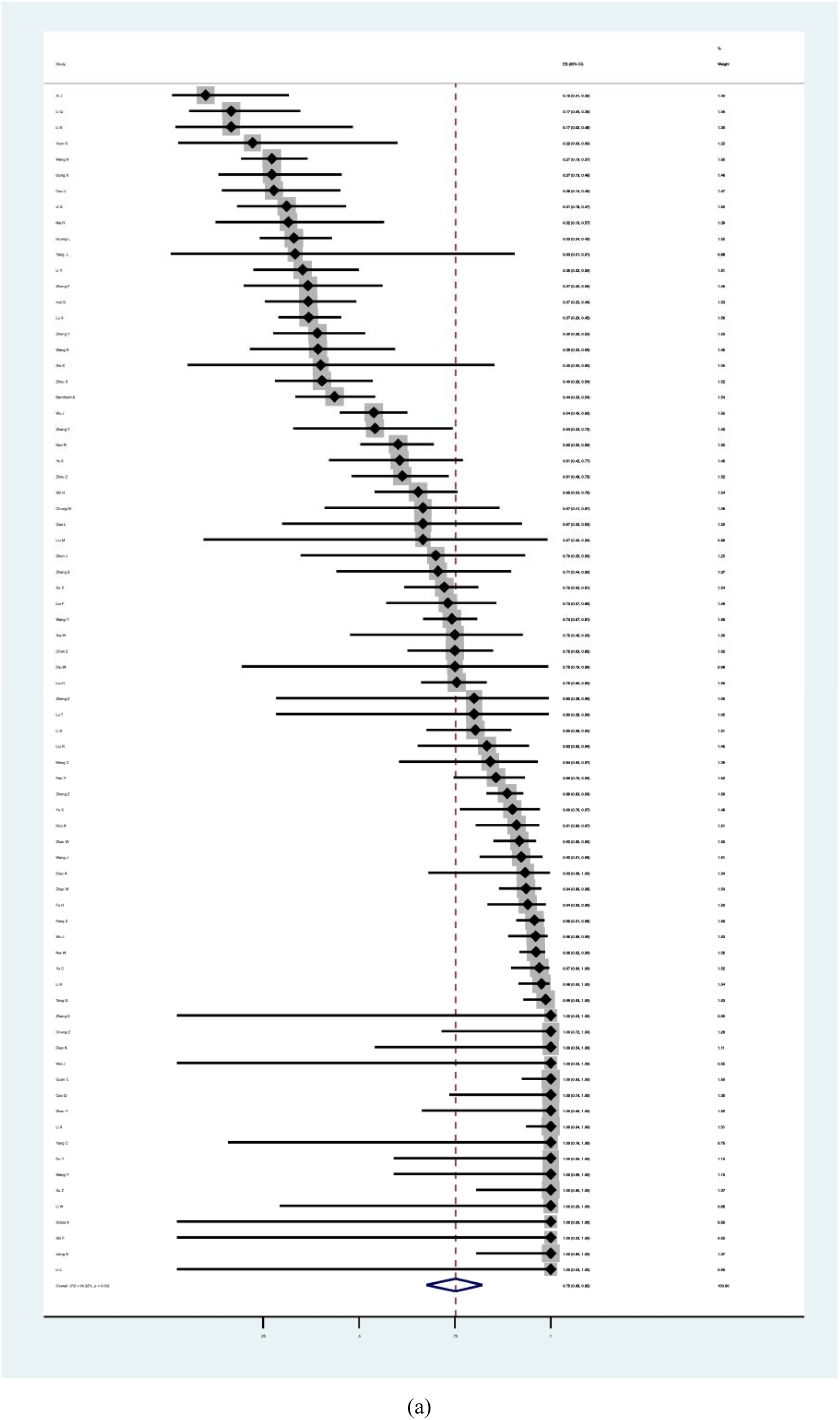
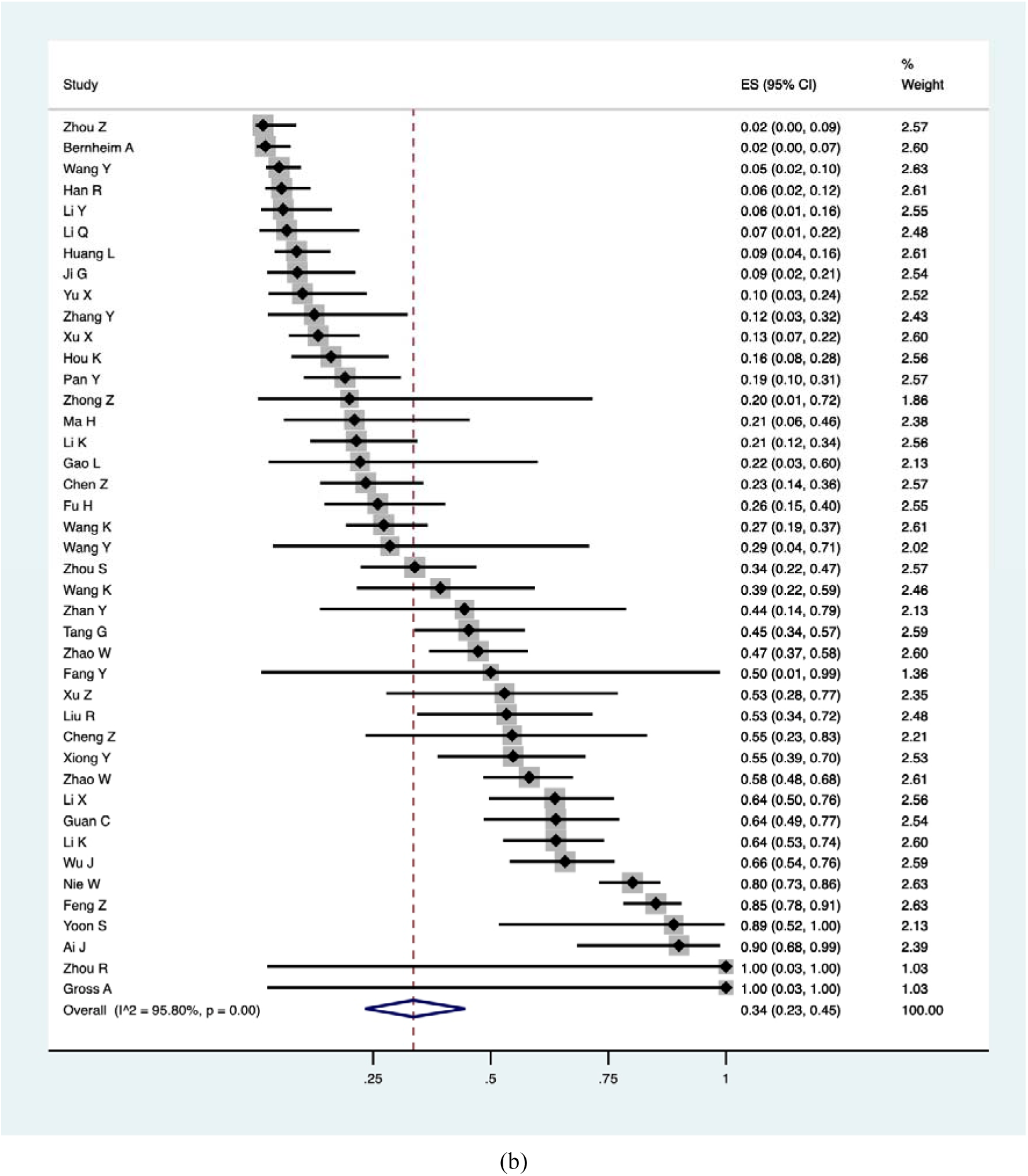
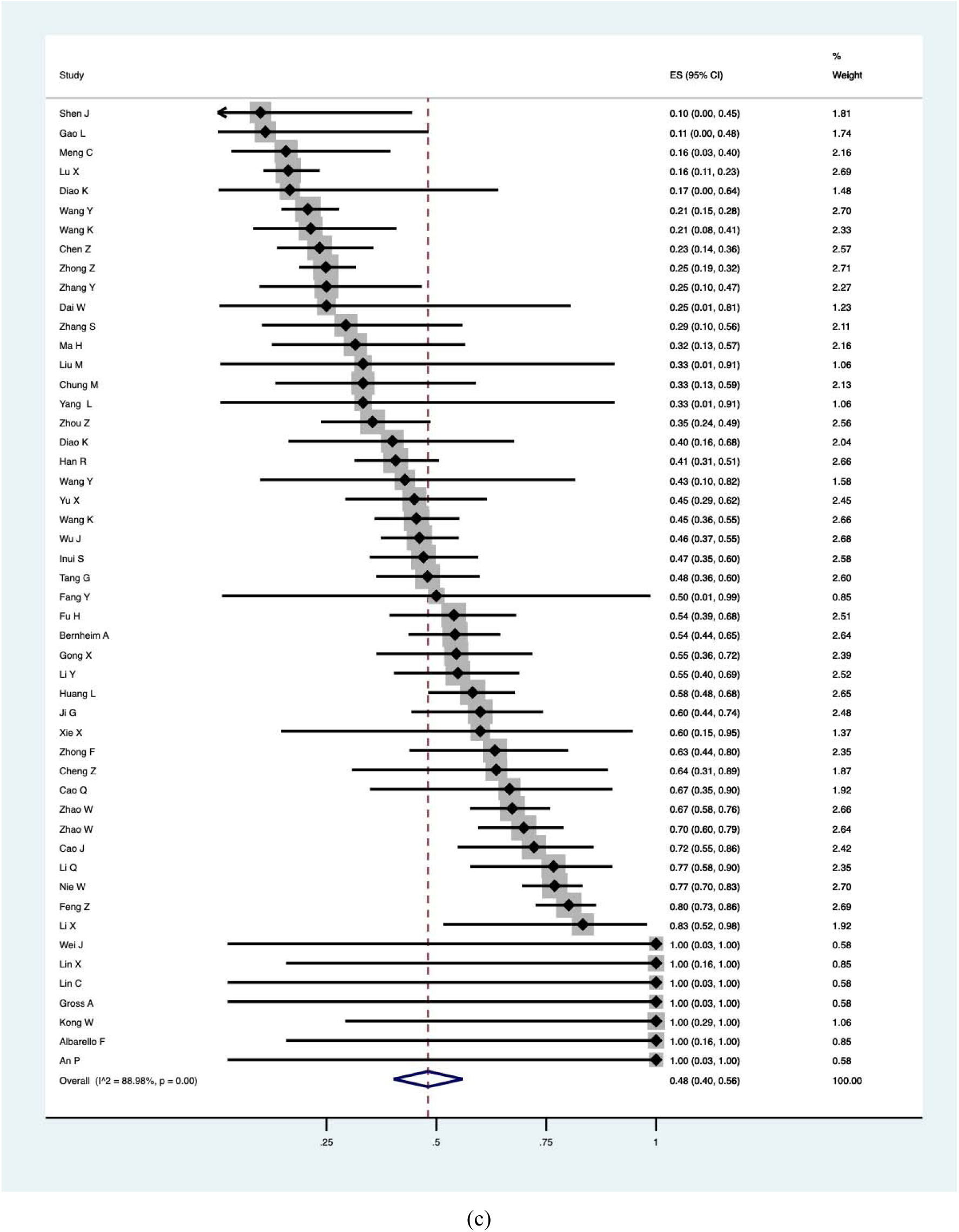
Meta-analyses of probability of lesion density: GGO (panel a); consolidation (panel b) and GGO with consolidation (panel c)

### Secondary outcomes

We conducted meta-analyses on the numbers of lobes affected, locations of lobes involved, and a total of 10 other secondary outcomes (*Table 1*). The quality of evidence for all secondary outcomes was low.

**Table 1.**
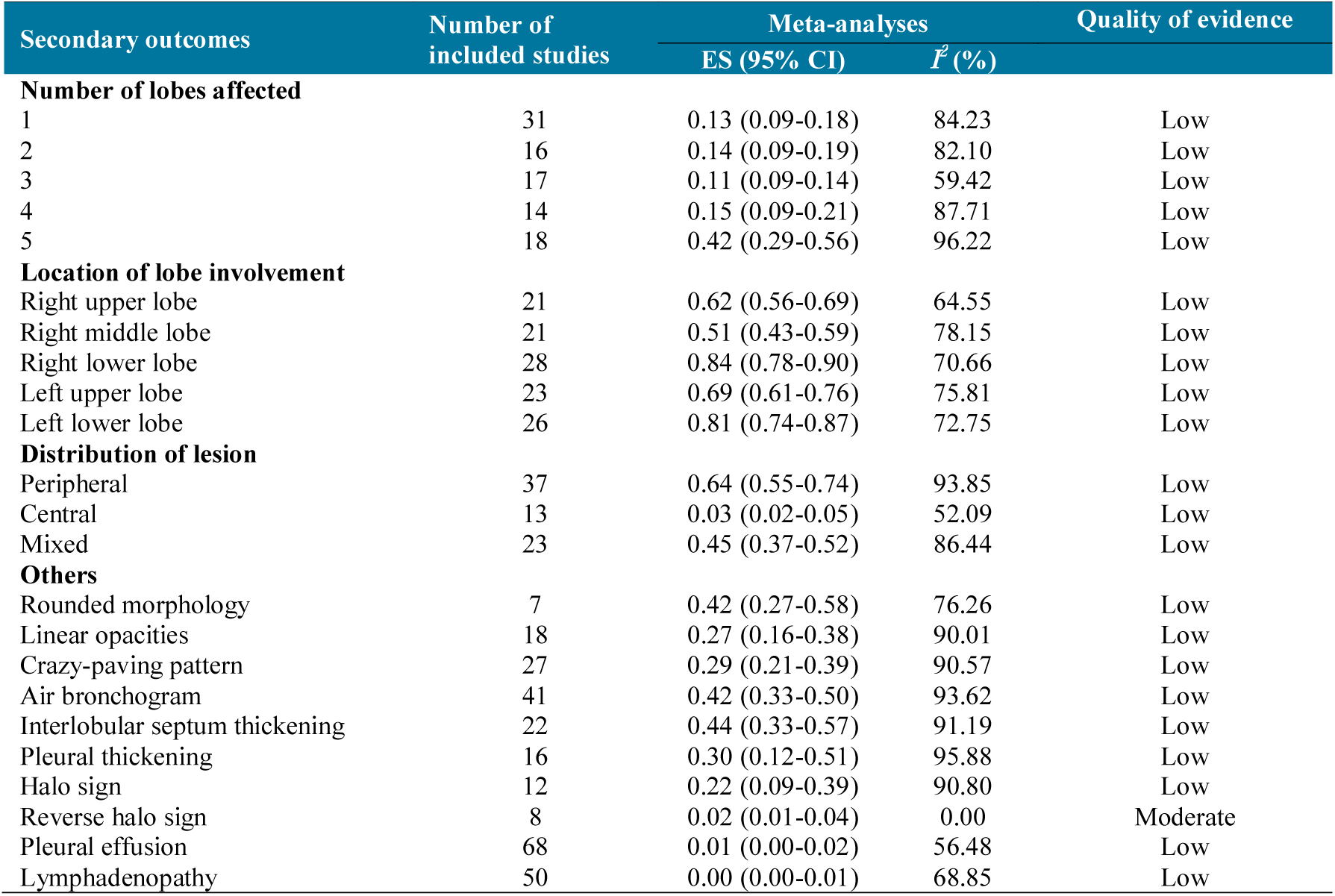
Meta-analyses of secondary outcomes

## Discussion

The sensitivity of chest CT imaging in patients with COVID-19 was 99% using RT-PCR results as reference. Therefore, CT scan can be useful in the diagnosis for people with COVID-19. However, the sensitivity of CT in children was 66%, which is much lower than it in general population. The most common imaging manifestation of patients infected with SARS-CoV-2 was GGO with bilateral peripheral distribution. The quality of evidence for almost all findings in our study was low.

Viral nucleic acid detection using RT-PCR is the gold standard in the diagnosis of COVID-19. However, some studies (22,23) reported that some patients had negative RT-PCR results, while their CT imaging features were abnormal. Several studies (24,25) have compared the diagnostic accuracy of chest CT scan and RT-PCR and found that the sensitivity of CT was higher than that of RT-PCR. A case series (24) with 1014 patients indicated that the sensitivity of chest CT scan for COVID-19 was 97%. Our study results also showed that the sensitivity of chest CT was 99%, which indicates that chest CT scan can effectively capture lung lesions in the early stage, especially in the epidemic areas. However, it is noteworthy that a small part of patients had normal CT imaging. Another systematic review (26) with 356 patients with COVID-19 also showed that 11.5% patients were diagnosed, while their CT imaging were normal, which revealed that CT examination cannot alone reliably fully exclude the diagnosis of COVID-19, notably in the early stage of infection.

Our meta-analysis showed that the most common type of imaging manifestation of patients with COVID-19 was GGO with bilateral peripheral distribution. This is consistent with a review (27), which also showed that the main imaging features in COVID-19 are GGO. The most commonly involved lobe was the right lower lobe, followed by the left lower lobe. Among the other signs, interlobular septum thickening was the most common, followed by rounded morphology and air bronchogram. Reverse halo sign, pleural effusion and lymphadenopathy were also rare. Although sensitivity of CT scan high, the specificity of CT in COVID-19 is limited, which need for differential diagnosis with other types of viral pneumonia (28).

Among the included studies, nine described the CT imaging features of children. The result indicated that COVID-19 tends to be mild in most children and the sensitivity of chest CT in children was only 66%. The role of CT in the diagnosis of COVID-19 in children is therefore limited. Some other studies (29,30) also indicated that most child patients had mild symptoms with atypical imaging findings. There is also so far no evidence to explicitly support the role of CT scan for the diagnosis of children with COVID-19. Considering that most children present only mild disease and the other risks in the process of using CT, such as radiation (31,32) and hospital-based transmission^5^, it is necessary to balance the advantages and disadvantages of CT use in the process of diagnosis in children with COVID-19.

Our review has several strengths. We performed a comprehensive search including databases and preprint servers and conducted meta-analyses on all main outcomes. The results of our study can thus help to better understand the role of CT imaging and the main CT manifestations in patients with COVID-19. However, this review also has some limitations: 1) though we conducted a systematic search, we only included articles published or posted in English and Chinese, which may introduce publication bias; 2) we only included case series and case reports, cases selection of included studies may introduce bias; 3) due to most of studies conducted in China, some cases may be overlapping between studies; and 4) there was large heterogeneity between included studies.

## Conclusion

In conclusion, this meta-analysis indicates that 98% of patients with COVID-19 present abnormal findings in their initial CT scan, suggesting that CT has the potential to be used as an assisting diagnostic tool. The most common imaging manifestation of patients with COVID-19 is GGO, and the probability of bilateral involvement was 84%. However, the quality of evidence was low across all outcomes. Studies with large sample size and clear reporting are needed in the future to guide the use of CT in the diagnosis and monitoring of patients with COVID-19.

## Data Availability

All data were shown as supplementary materials in the PDF.

## Author contributions

(I) Conception and design: Y Chen, X Wang and J Tian; (II) Administrative support: J Tian and X Wang; (III) Provision of study materials or patients: M Lv, M and Wang; (IV) Collection and assembly of data: N Yang, X Luo, W Li, X Chen, Y Liu, M Ren, X Zhang and L Wang; (V) Data analysis and interpretation: M Lv, M Wang and Y Ma; (VI) Manuscript writing: All authors; (VII) Final approval of manuscript: All authors.

## Acknowledgements

We thank Janne Estill, Institute of Global Health of University of Geneva for providing guidance and comments for our review. We thank all the authors for their wonderful collaboration.

## Footnote

### Ethical Statement

The authors are accountable for all aspects of the work in ensuring that questions related to the accuracy or integrity of any part of the work are appropriately investigated and resolved.

## Supplementary Material 1-Search strategy

### PubMed (n=138)

#1 “COVID-19”[Supplementary Concept]

#2 “Severe Acute Respiratory Syndrome Coronavirus 2”[Supplementary Concept] #3 “COVID-19”[Title/Abstract]

#4 “SARS-COV-2”[Title/Abstract]

#5 “Novel coronavirus” [Title/Abstract]

#6 “2019-novel coronavirus” [Title/Abstract]

#7 “coronavirus disease-19” [Title/Abstract]

#8 “coronavirus disease 2019” [Title/Abstract]

#9 “COVID19” [Title/Abstract]

#10 “Novel CoV” [Title/Abstract]

#11 “2019-nCoV” [Title/Abstract]

#12 “2019-CoV” [Title/Abstract]

#13 “Wuhan-Cov” [Title/Abstract]

#14 “Wuhan Coronavirus” [Title/Abstract]

#15 “Wuhan seafood market pneumonia virus” [Title/Abstract]

#16 #1-#15/ OR

#17 “Radiography, Thoracic”[Mesh]

#18 “computed tomography”[Title/Abstract]

#19 radiograph*[Title/Abstract]

#20 imagin*[Title/Abstract]

#21 #17-#20/ OR

#22 #16 AND #21

### EMBASE (n=72)

#1 ‘COVID-19’:ab,ti

#2 ‘SARS-COV-2’:ab,ti

#3 ‘novel coronavirus’:ab,ti

#4 ‘2019-novel coronavirus’:ab,ti

#5‘coronavirus disease-19’:ab,ti

#6 ‘coronavirus disease 2019’:ab,ti

#7 ‘COVID19’:ab,ti

#8 ‘novel cov’:ab,ti

#9 ‘2019-ncov’:ab,ti

#10 ‘2019-cov’:ab,ti

#11 ‘wuhan-cov’:ab,ti

#12 ‘wuhan coronavirus’:ab,ti

#13 ‘wuhan seafood market pneumonia virus’:ab,ti

#14 #1-#13/ OR

#15 thorax radiography’/exp

#16 ‘computed tomography’:ab,ti

#17 radiograph*:ab,ti

#18 Imagin*:ab,ti

#19 #15-#18/ OR

#20 #14 AND #19

### Cochrane Library (n=5)

#1 “COVID-19”:ti,ab,kw

#2 “SARS-COV-2”:ti,ab,kw

#3 “Novel coronavirus”:ti,ab,kw

#4 “2019-novel coronavirus” :ti,ab,kw

#5 “Novel CoV” :ti,ab,kw

#6 “2019-nCoV” :ti,ab,kw

#7 “2019-CoV” :ti,ab,kw

#8 “coronavirus disease-19” :ti,ab,kw

#9 “coronavirus disease 2019” :ti,ab,kw

#10 “COVID19” :ti,ab,kw

#11 “Wuhan-Cov” :ti,ab,kw

#12 “Wuhan Coronavirus” :ti,ab,kw

#13 “Wuhan seafood market pneumonia virus” :ti,ab,kw

#14 #1-#13/ OR

#15 MeSH descriptor: [Radiography, Thoracic] explode all trees

#16 “computed tomography”:ti,ab,kw

#17 radiograph*:ti,ab,kw

#18 imagin*:ti,ab,kw

#19 #15-#18/ OR

#20 #14 AND #19

### Web of Science (n=27)

#1 TOPIC: “COVID-19”

#2 TOPIC: “SARS-COV-2”

#3 TOPIC: “Novel coronavirus”

#4 TOPIC: “2019-novel coronavirus”

#5 TOPIC: “coronavirus disease-19” [Title/Abstract]

#6 TOPIC: “coronavirus disease 2019” [Title/Abstract]

#7 TOPIC: “COVID19” [Title/Abstract]

#8 TOPIC: “Novel CoV”

#9 TOPIC: “2019-nCoV”

#10 TOPIC: “2019-CoV”

#11 TOPIC: “Wuhan-Cov”

#12 TOPIC: “Wuhan Coronavirus”

#13 TOPIC: “Wuhan seafood market pneumonia virus”

#14 #1-#13/ OR

#15 TOPIC: “computed tomography”

#16 TOPIC: “radiograph*”

#17 TOPIC: “imagin*”

#18 #15-#17/ OR

#19 #14 AND #18

### CNKI (n=52)

#1 “新型冠状病毒”[主题]

#2 “2019-nCoV”[主题]

#3 “2019-CoV”[主题]

#4 “武汉冠状病毒”[主题]

#5 “COVID-19” [主题]

#6 “COVID 19”[主题]

#7 “SARS-CoV-2” [主题]

#8 #1-#7/ OR

#9 “CT 扫描”[主题]

#10 “CAT 扫描” [主题]

#11 “电子束计算机断层摄影术”[主题]

#12 “断层摄影术”[主题]

#13 “计算机断层摄影术” [主题]

#14 #9-#13/ OR

#15 #8 AND #14

### CBM (n=163)

#1 “新型冠状病毒” [常用字段:智能]

#2 “2019-nCoV”[常用字段:智能]

#3 “2019-CoV”[常用字段:智能]

#4 “武汉冠状病毒”[常用字段:智能]

#5 “COVID-19” [常用字段:智能]

#6 “COVID 19”[常用字段:智能]

#7 “SARS-CoV-2” [常用字段:智能]

#8 #1-#7/ OR

#9 “CT 扫描&#x201D;[常用字段:智能]

#10 “CAT 扫描” [常用字段:智能]

#11 “电子束计算机断层摄影术”[常用字段:智能]

#12 “断层摄影术”[常用字段:智能]

#13 “计算机断层摄影术” [常用字段:智能]

#14 #9-#13/ OR

#15 #8 AND #14

### WangFang (n=36)

#1 “新型冠状病毒”[主题]

#2 “2019-nCoV”[主题]

#3 “2019-CoV”[主题]

#4 “武汉冠状病毒”[主题]

#5 “COVID-19” [主题]

#6 “COVID 19”[主题]

#7 “SARS-CoV-2” [主题]

#8 #1-#7/ OR

#9 “CT 扫描”[主题]

#10 “CAT 扫描” [主题]

#11 “电子束计算机断层摄影术”[主题]

#12 “断层摄影术”[主题]

#13 “计算机断层摄影术” [主题]

#14 #9-#13/ OR

#15 #8 AND #14

---

**Supplementary Material 3.**
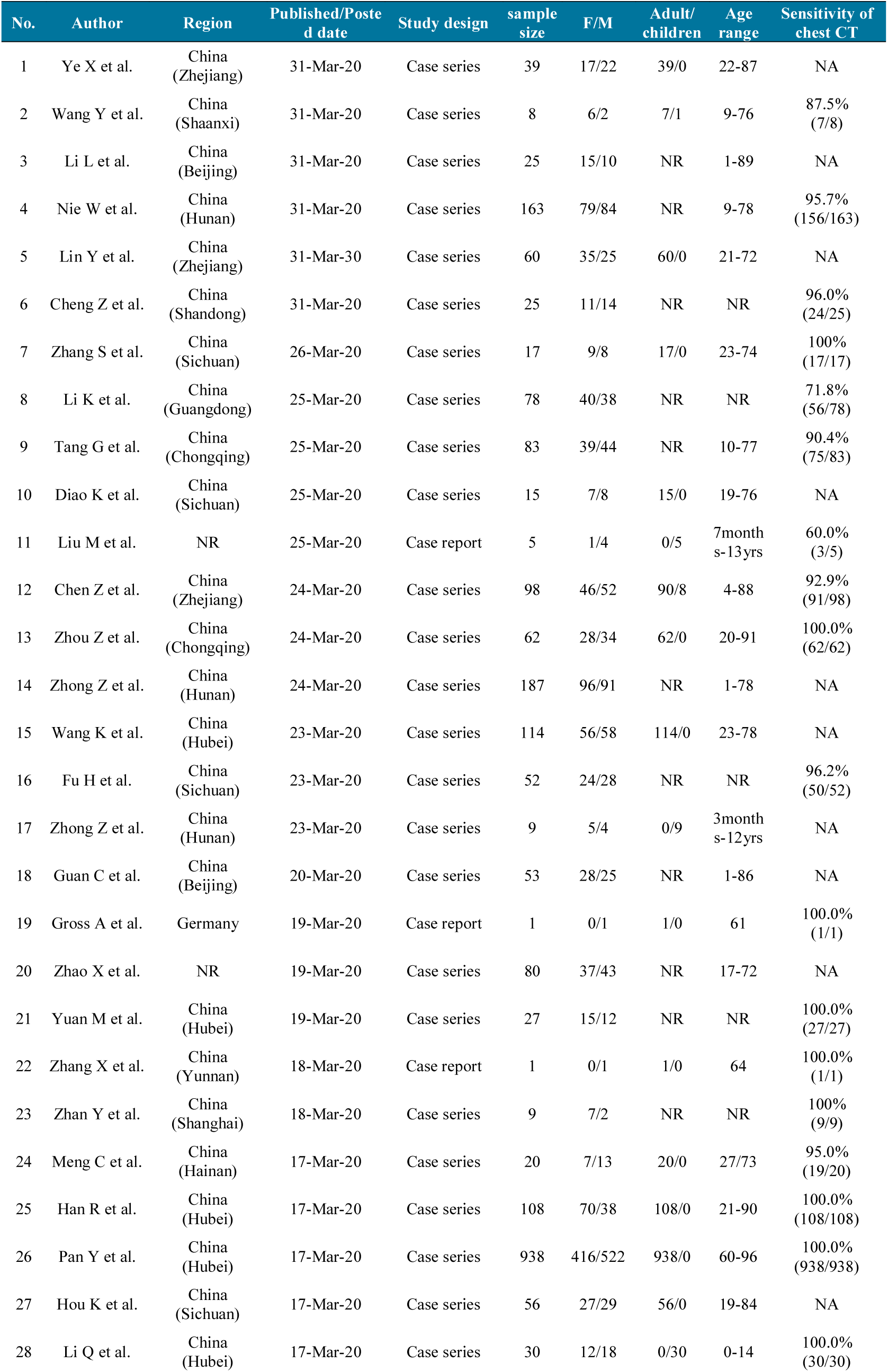
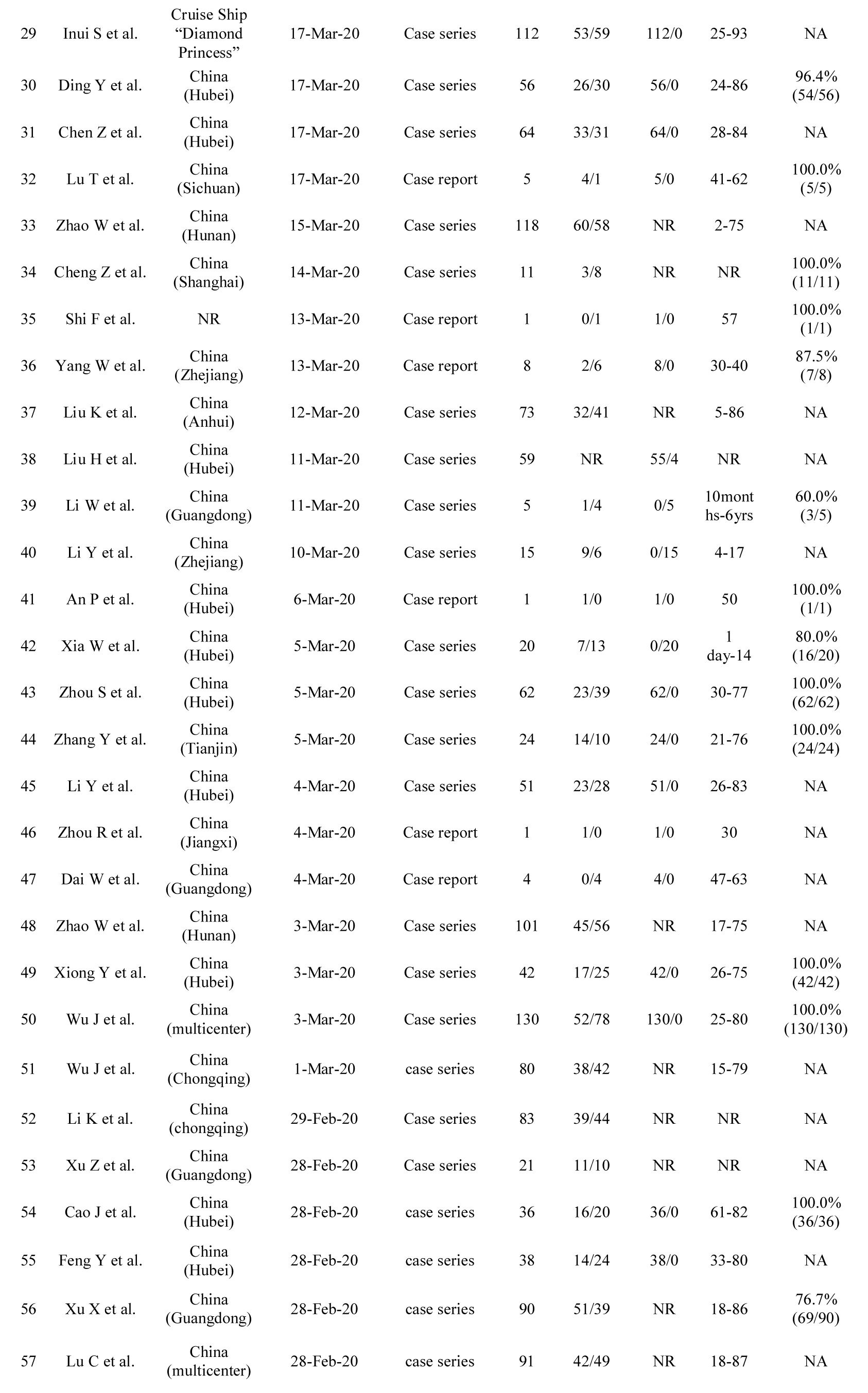
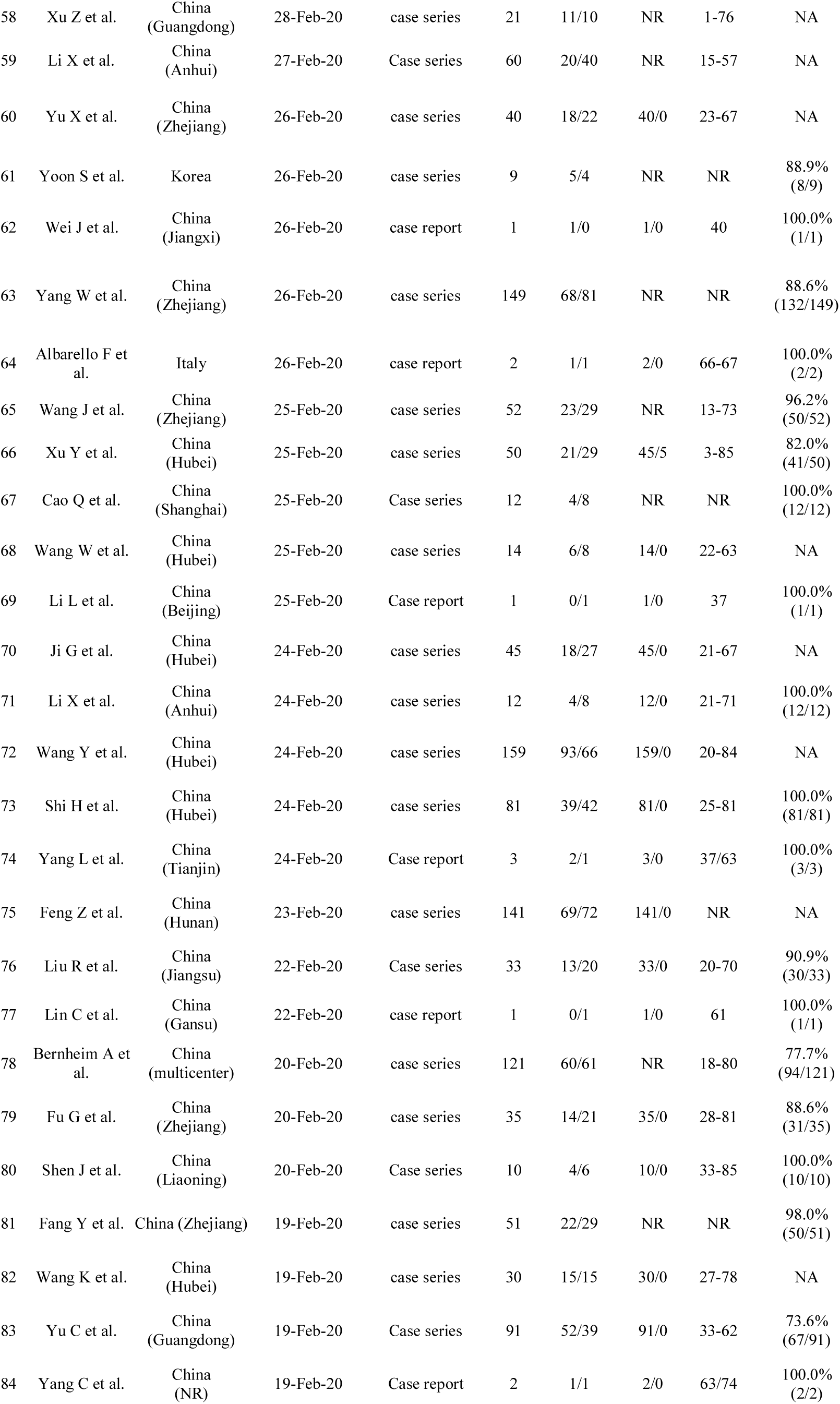
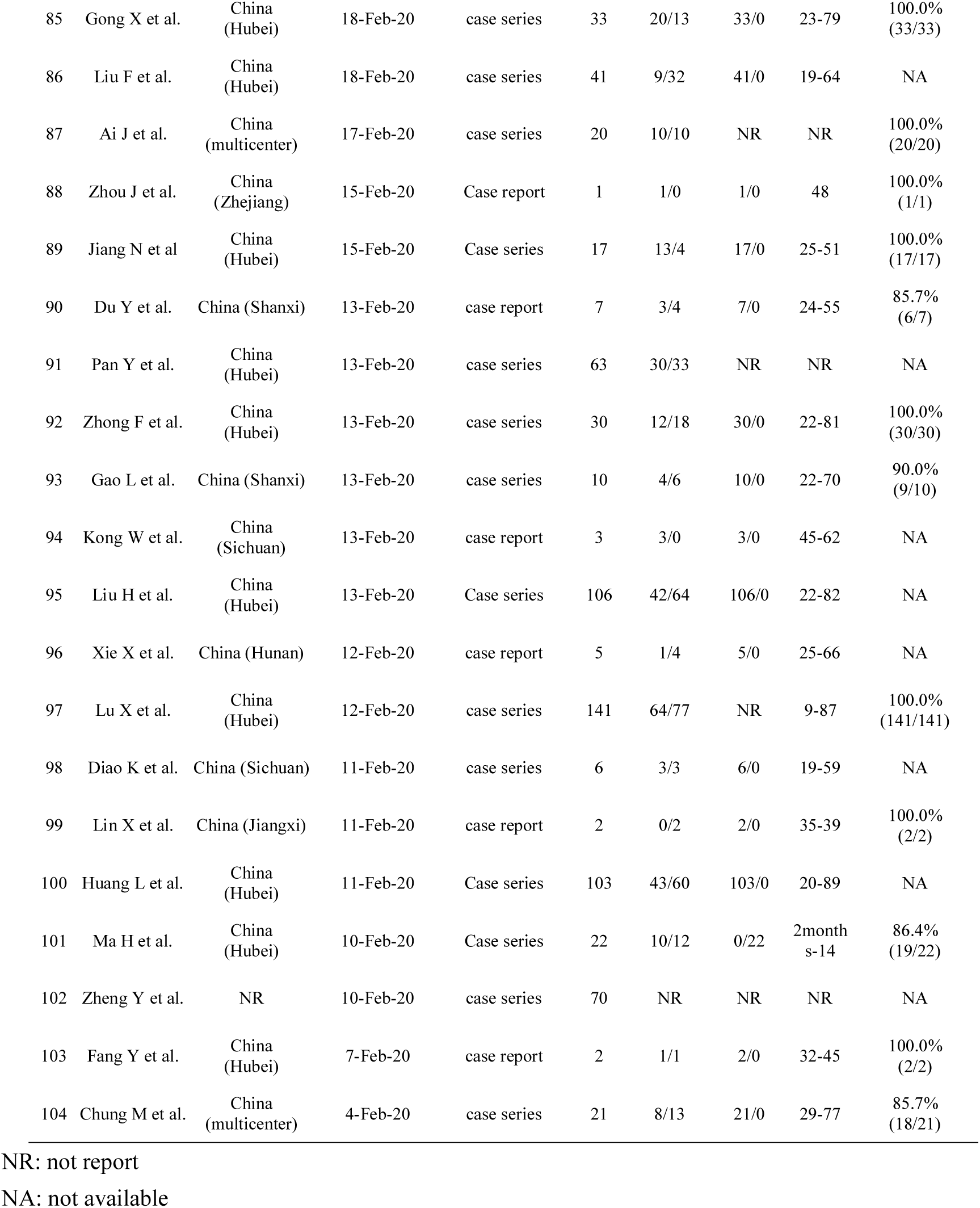
The characteristics of the included studies

**Supplementary Table 4.**
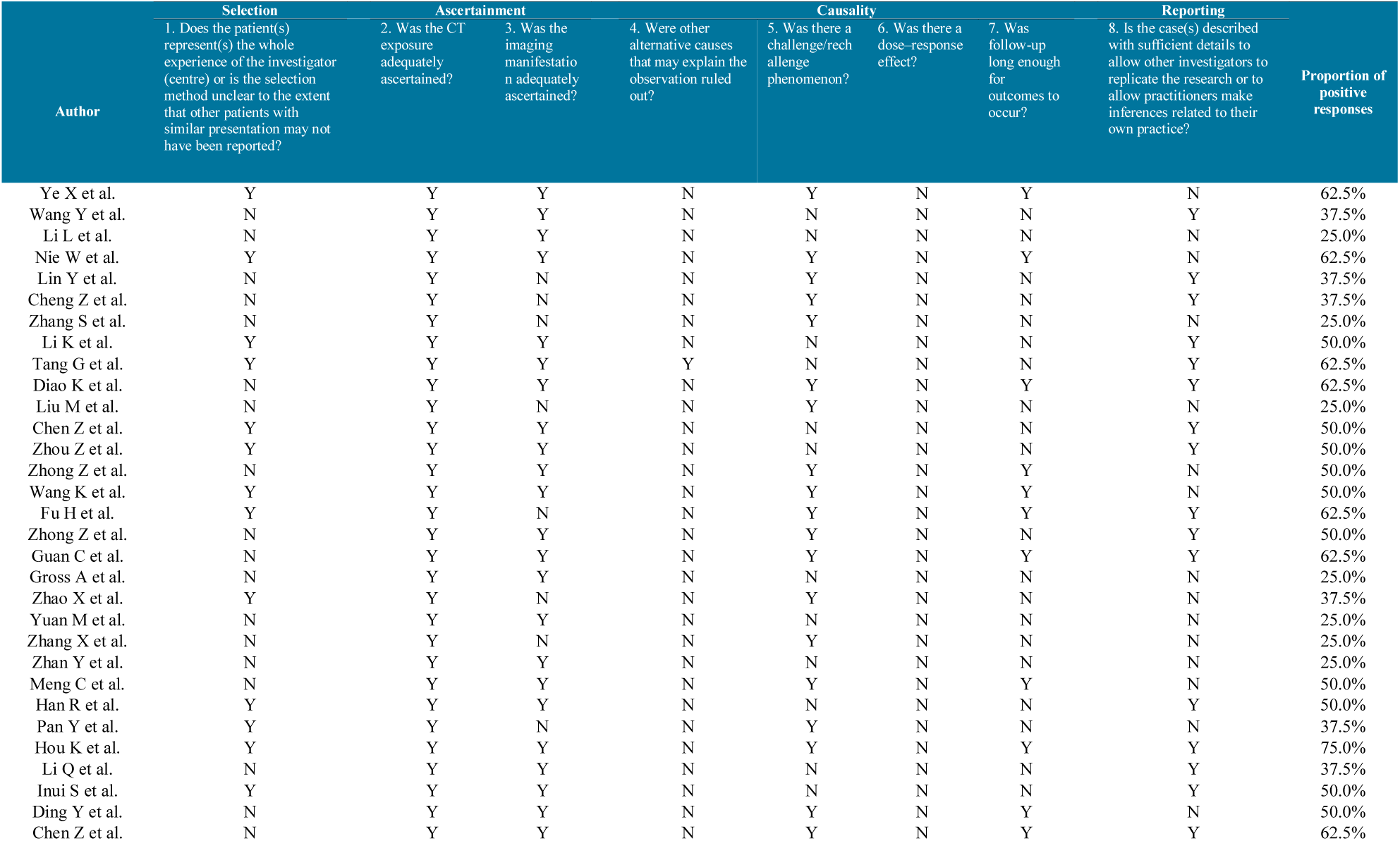
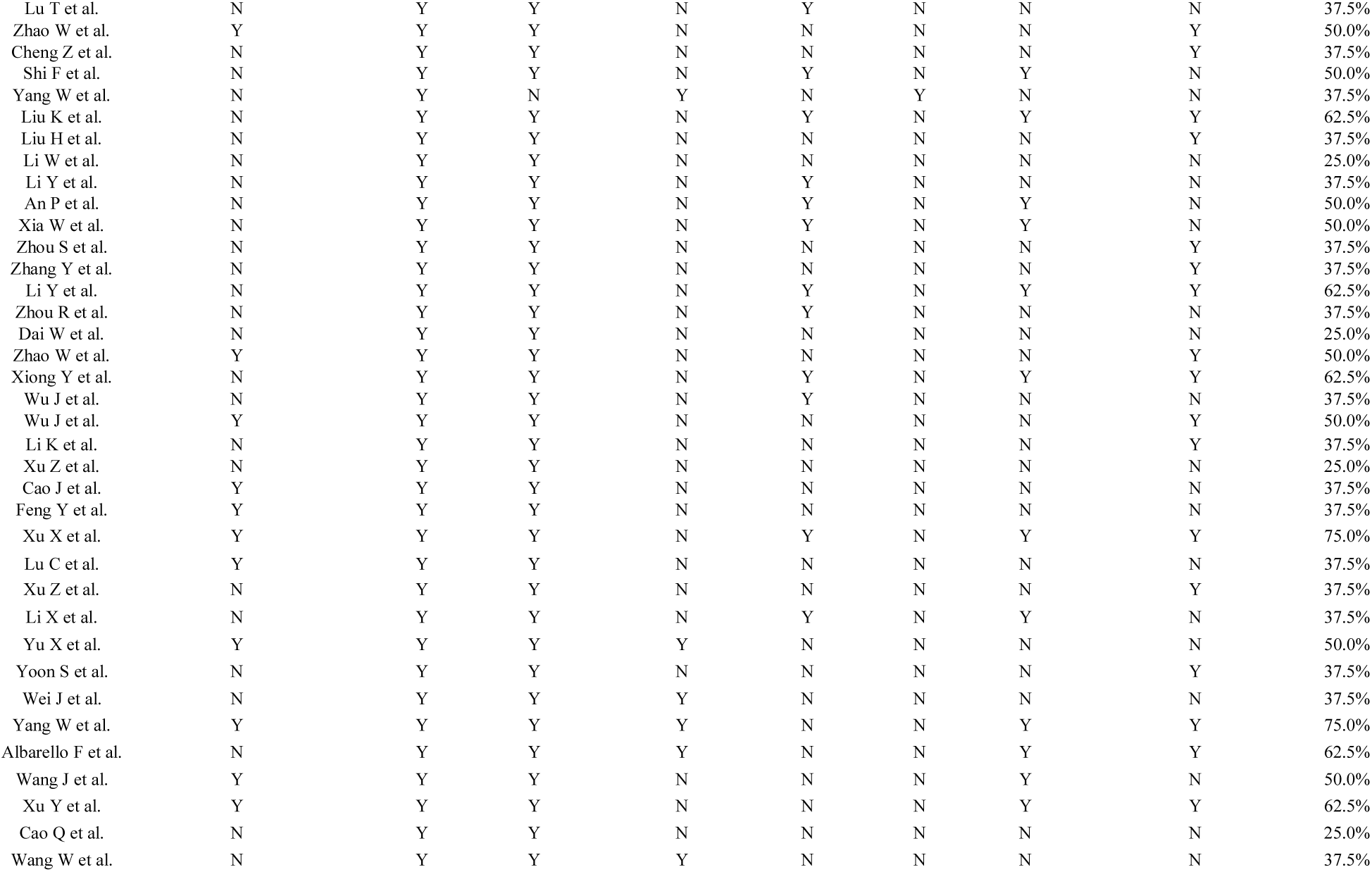
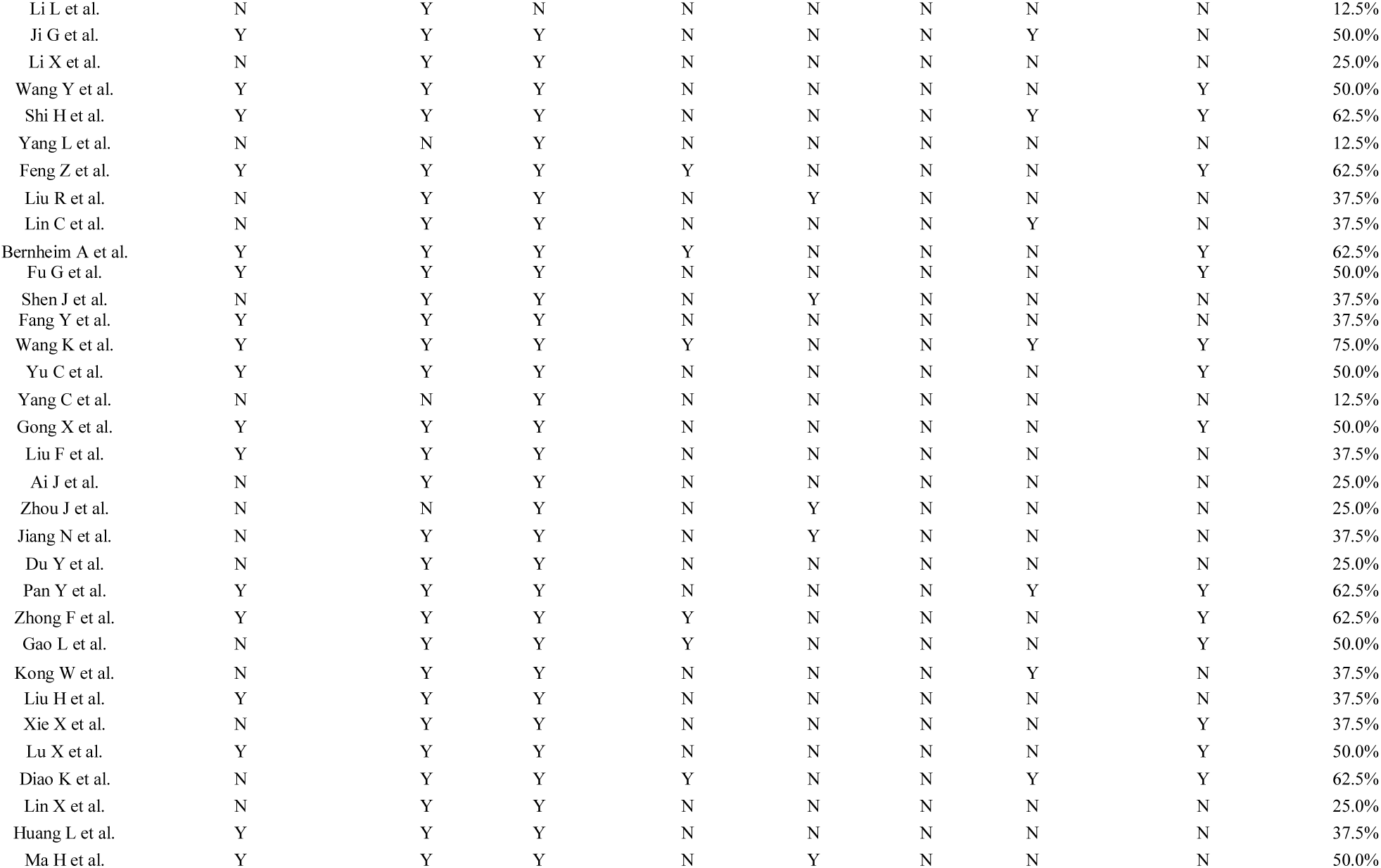
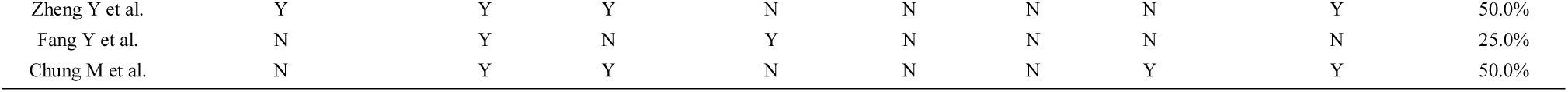
Risk of bias of the included studies

Supplementary Material 4-Risk of bias of the included studies
| Author | Selection | Ascertainment |  | Causality |  |  |  | Reporting | Proportion of positive responses |
| --- | --- | --- | --- | --- | --- | --- | --- | --- | --- |
|  | 1. Does the patient(s) represent(s) the whole experience of the investigator (centre) or is the selection method unclear to the extent that other patients with similar presentation may not have been reported? | 2. Was the CT exposure adequately ascertained? | 3. Was the imaging manifestation adequately ascertained? | 4. Were other alternative causes that may explain the observation ruled out? | 5. Was there a challenge/rechallenge phenomenon? | 6. Was there a dose-response effect? | 7. Was follow-up long enough for outcomes to occur? | 8. Is the case(s) described with sufficient details to allow other investigators to replicate the research or to allow practitioners make inferences related to their own practice? |  |
| Ye X et al. | Y | Y | Y | N | Y | N | Y | N | 62.5% |
| Wang Y et al. | N | Y | Y | N | N | N | N | Y | 37.5% |
| Li L et al. | N | Y | Y | N | N | N | N | N | 25.0% |
| Nie W et al. | Y | Y | Y | N | Y | N | Y | N | 62.5% |
| Lin Y et al. | N | Y | N | N | Y | N | N | Y | 37.5% |
| Cheng Z et al. | N | Y | N | N | Y | N | N | Y | 37.5% |
| Zhang S et al. | N | Y | N | N | Y | N | N | N | 25.0% |
| Li K et al. | Y | Y | Y | N | N | N | N | Y | 50.0% |
| Tang G et al. | Y | Y | Y | Y | N | N | N | Y | 62.5% |
| Diao K et al. | N | Y | Y | N | Y | N | Y | Y | 62.5% |
| Liu M et al. | N | Y | N | N | Y | N | N | N | 25.0% |
| Chen Z et al. | Y | Y | Y | N | N | N | N | Y | 50.0% |
| Zhou Z et al. | Y | Y | Y | N | N | N | N | Y | 50.0% |
| Zhong Z et al. | N | Y | Y | N | Y | N | Y | N | 50.0% |
| Wang K et al. | Y | Y | Y | N | Y | N | Y | N | 50.0% |
| Fu H et al. | Y | Y | N | N | Y | N | Y | Y | 62.5% |
| Zhong Z et al. | N | Y | Y | N | Y | N | N | Y | 50.0% |
| Guan C et al. | N | Y | Y | N | Y | N | Y | Y | 62.5% |
| Gross A et al. | N | Y | Y | N | N | N | N | N | 25.0% |
| Zhao X et al. | Y | Y | N | N | Y | N | N | N | 37.5% |
| Yuan M et al. | N | Y | Y | N | N | N | N | N | 25.0% |
| Zhang X et al. | N | Y | N | N | Y | N | N | N | 25.0% |
| Zhan Y et al. | N | Y | Y | N | N | N | N | N | 25.0% |
| Meng C et al. | N | Y | Y | N | Y | N | Y | N | 50.0% |
| Han R et al. | Y | Y | Y | N | N | N | N | Y | 50.0% |
| Pan Y et al. | Y | Y | N | N | Y | N | N | N | 37.5% |
| Hou K et al. | Y | Y | Y | N | Y | N | Y | Y | 75.0% |
| Li Q et al. | N | Y | Y | N | N | N | N | Y | 37.5% |
| Inui S et al. | Y | Y | Y | N | N | N | N | Y | 50.0% |
| Ding Y et al. | N | Y | Y | N | Y | N | Y | N | 50.0% |
| Chen Z et al. | N | Y | Y | N | Y | N | Y | Y | 62.5% |
| Lu T et al. | N | Y | Y | N | Y | N | N | N | 37.5% |
| Zhao W et al. | Y | Y | Y | N | N | N | N | Y | 50.0% |
| Cheng Z et al. | N | Y | Y | N | N | N | N | Y | 37.5% |
| Shi F et al. | N | Y | Y | N | Y | N | Y | N | 50.0% |
| Yang W et al. | N | Y | N | Y | N | Y | N | N | 37.5% |
| Liu K et al. | N | Y | Y | N | Y | N | Y | Y | 62.5% |
| Liu H et al. | N | Y | Y | N | N | N | N | Y | 37.5% |
| Li W et al. | N | Y | Y | N | N | N | N | N | 25.0% |
| Li Y et al. | N | Y | Y | N | Y | N | N | N | 37.5% |
| An P et al. | N | Y | Y | N | Y | N | Y | N | 50.0% |
| Xia W et al. | N | Y | Y | N | Y | N | Y | N | 50.0% |
| Zhou S et al. | N | Y | Y | N | N | N | N | Y | 37.5% |
| Zhang Y et al. | N | Y | Y | N | N | N | N | Y | 37.5% |
| Li Y et al. | N | Y | Y | N | Y | N | Y | Y | 62.5% |
| Zhou R et al. | N | Y | Y | N | Y | N | N | N | 37.5% |
| Dai W et al. | N | Y | Y | N | N | N | N | N | 25.0% |
| Zhao W et al. | Y | Y | Y | N | N | N | N | Y | 50.0% |
| Xiong Y et al. | N | Y | Y | N | Y | N | Y | Y | 62.5% |
| Wu J et al. | N | Y | Y | N | Y | N | N | N | 37.5% |
| Wu J et al. | Y | Y | Y | N | N | N | N | Y | 50.0% |
| Li K et al. | N | Y | Y | N | N | N | N | Y | 37.5% |
| Xu Z et al. | N | Y | Y | N | N | N | N | N | 25.0% |
| Cao J et al. | Y | Y | Y | N | N | N | N | N | 37.5% |
| Feng Y et al. | Y | Y | Y | N | N | N | N | N | 37.5% |
| Xu X et al. | Y | Y | Y | N | Y | N | Y | Y | 75.0% |
| Lu C et al. | Y | Y | Y | N | N | N | N | N | 37.5% |
| Xu Z et al. | N | Y | Y | N | N | N | N | Y | 37.5% |
| Li X et al. | N | Y | Y | N | Y | N | Y | N | 37.5% |
| Yu X et al. | Y | Y | Y | Y | N | N | N | N | 50.0% |
| Yoon S et al. | N | Y | Y | N | N | N | N | Y | 37.5% |
| Wei J et al. | N | Y | Y | Y | N | N | N | N | 37.5% |
| Yang W et al. | Y | Y | Y | Y | N | N | Y | Y | 75.0% |
| Albarello F et al. | N | Y | Y | Y | N | N | Y | Y | 62.5% |
| Wang J et al. | Y | Y | Y | N | N | N | Y | N | 50.0% |
| Xu Y et al. | Y | Y | Y | N | N | N | Y | Y | 62.5% |
| Cao Q et al. | N | Y | Y | N | N | N | N | N | 25.0% |
| Wang W et al. | N | Y | Y | Y | N | N | N | N | 37.5% |
| Li L et al. | N | Y | N | N | N | N | N | N | 12.5% |
| Ji G et al. | Y | Y | Y | N | N | N | Y | N | 50.0% |
| Li X et al. | N | Y | Y | N | N | N | N | N | 25.0% |
| Wang Y et al. | Y | Y | Y | N | N | N | N | Y | 50.0% |
| Shi H et al. | Y | Y | Y | N | N | N | Y | Y | 62.5% |
| Yang L et al. | N | N | Y | N | N | N | N | N | 12.5% |
| Feng Z et al. | Y | Y | Y | Y | N | N | N | Y | 62.5% |
| Liu R et al. | N | Y | Y | N | Y | N | N | N | 37.5% |
| Lin C et al. | N | Y | Y | N | N | N | Y | N | 37.5% |
| Bernheim A et al. | Y | Y | Y | Y | N | N | N | Y | 62.5% |
| Fu G et al. | Y | Y | Y | N | N | N | N | Y | 50.0% |
| Shen J et al. | N | Y | Y | N | Y | N | N | N | 37.5% |
| Fang Y et al. | Y | Y | Y | N | N | N | N | N | 37.5% |
| Wang K et al. | Y | Y | Y | Y | N | N | Y | Y | 75.0% |
| Yu C et al. | Y | Y | Y | N | N | N | N | Y | 50.0% |
| Yang C et al. | N | N | Y | N | N | N | N | N | 12.5% |
| Gong X et al. | Y | Y | Y | N | N | N | N | Y | 50.0% |
| Liu F et al. | Y | Y | Y | N | N | N | N | N | 37.5% |
| Ai J et al. | N | Y | Y | N | N | N | N | N | 25.0% |
| Zhou J et al. | N | N | Y | N | Y | N | N | N | 25.0% |
| Jiang N et al. | N | Y | Y | N | Y | N | N | N | 37.5% |
| Du Y et al. | N | Y | Y | N | N | N | N | N | 25.0% |
| Pan Y et al. | Y | Y | Y | N | N | N | Y | Y | 62.5% |
| Zhong F et al. | Y | Y | Y | Y | N | N | N | Y | 62.5% |
| Gao L et al. | N | Y | Y | Y | N | N | N | Y | 50.0% |
| Kong W et al. | N | Y | Y | N | N | N | Y | N | 37.5% |
| Liu H et al. | Y | Y | Y | N | N | N | N | N | 37.5% |
| Xie X et al. | N | Y | Y | N | N | N | N | Y | 37.5% |
| Lu X et al. | Y | Y | Y | N | N | N | N | Y | 50.0% |
| Diao K et al. | N | Y | Y | Y | N | N | Y | Y | 62.5% |
| Lin X et al. | N | Y | Y | N | N | N | N | N | 25.0% |
| Huang L et al. | Y | Y | Y | N | N | N | N | N | 37.5% |
| Ma H et al. | Y | Y | Y | N | Y | N | N | N | 50.0% |
| Zheng Y et al. | Y | Y | Y | N | N | N | N | Y | 50.0% |
| Fang Y et al. | N | Y | N | Y | N | N | N | N | 25.0% |
| Chung M et al. | N | Y | Y | N | N | N | Y | Y | 50.0% |

